# Early assessment of epidemiological trends associated with SARS-CoV-2 variants of concern in Germany

**DOI:** 10.1101/2021.02.16.21251803

**Authors:** Timo Mitze, Johannes Rode

## Abstract

Growing evidence on higher transmissibility of novel variants of the SARS-CoV-2 coronavirus is raising alarm in many countries. We provide near-time estimates of the statistical association between reported cases of SARS-CoV-2 variants of concern (VOC) and epidemiological indicators at the local area level in Germany. Our findings indicate that the 7-day incidence rates in regions with confirmed VOC cases increased by up to 35%, on average, after VOC reporting compared to regions without confirmed cases by February 4. The hospitalization rate for COVID-19 patients in intensive care increased by up to 40%, but only for regions with most reported VOC cases. Both indicators further show a clear upward trend in regions with reported VOC cases vis-à-vis those without cases.

---

Newly emerging variants of concern (VOC) of the SARS-CoV-2 coronavirus, including the British (B.1.1.7.), South African (B.1.351) and Brazilian (P.1.) mutations, constitute a significant threat for public health systems around the globe. First epidemiological evidence points to an increased transmissibility of the British and South African variants compared to previously circulating strains of SARS-CoV-2 of about 50-70% [1, 2, 3, 4]. There is also evidence for potentially higher death rates associated with the British variant [5] and an enhanced risk of international disease transmission [6]. By mid-January 2021, all three VOC were confirmed in Germany.

The main goal of this study is to provide near-time estimates of the epidemiological trends associated with the reporting of these novel strains for two key indicators at the local area level in Germany: i) the 7-day incidence rate, i.e., the number of newly reported SARS-CoV-2 infections in the last seven days per 100,000 population, and ii) the hospitalization rate, i.e., the number of hospitalized COVID-19 patients in intensive care per 100,000 population. While the 7-day incidence rate is the main indicator used for disease surveillance and public health decisions in Germany, the hospitalization rate is used here as an important stress indicator for local health care systems.

A severe challenge for this endeavor is that genomic data on VOC spread is still very limited in Germany with prior analyses being largely based on ad-hoc laboratory sampling procedures [7]. As a solution to this problem, we use an event database that collects information on confirmed VOC cases by genome sequencing of the three most concerning VOC together with their reporting dates in a public crowd-sourcing project [8]. Out of all clearly identifiable cases until February 4, 88.2% dated from the British B.1.1.7., 11.5% from the B.1.351 South African and 0.3% from the P.1 Brazilian virus mutation. Case documentation is based on newspaper and further public health reports.

By February 4, in 204 out of 401 NUTS-3 regions at least one case of a SARS-CoV-2 infection from one of the three VOC (B.1.1.7, B.1.351, P.1) had been confirmed and the number of affected regions had been growing exponentially by the end of January (see Panel A of Figure 1). Panel C of Figure 1 displays the spatial distribution of cumulative VOC cases across all three variants. The map reveals that Flensburg and a cluster of three NUTS-3 regions (Cologne, Leverkusen and Düren) in North-Rhine Westphalia (NRW) are among the top 5% percentile of most affected regions in Germany.

A first inspection of incidence rate development in these regions nourishes concerns that virus mutations significantly drive the local infection dynamics. In Flensburg, a 90,000-inhabitant city in Schleswig-Holstein, the 7-day incidence rate drastically increased in January 2021 relative to the average development in Schleswig-Holstein and Germany (see Panel B of Figure 1). According to local health authorities, new infections mainly happened at illegal parties, i.e., against prevailing lockdown rules, on New Year’s Eve. On January 24, 2021, for several SARS-CoV-2 infections related to these parties, but also unrelated instances, the British B.1.1.7. VOC was confirmed. By February 4, the number of VOC cases had grown to 146 in Flensburg. In the three NRW cities, first VOC cases were reported between January 12 and January 23. While for Düren only the British B.1.1.7. variant was identified through genome sequencing, Cologne and Leverkusen reported both the British B.1.1.7. and the South African B.1.351. variants.

**Figure 1.**
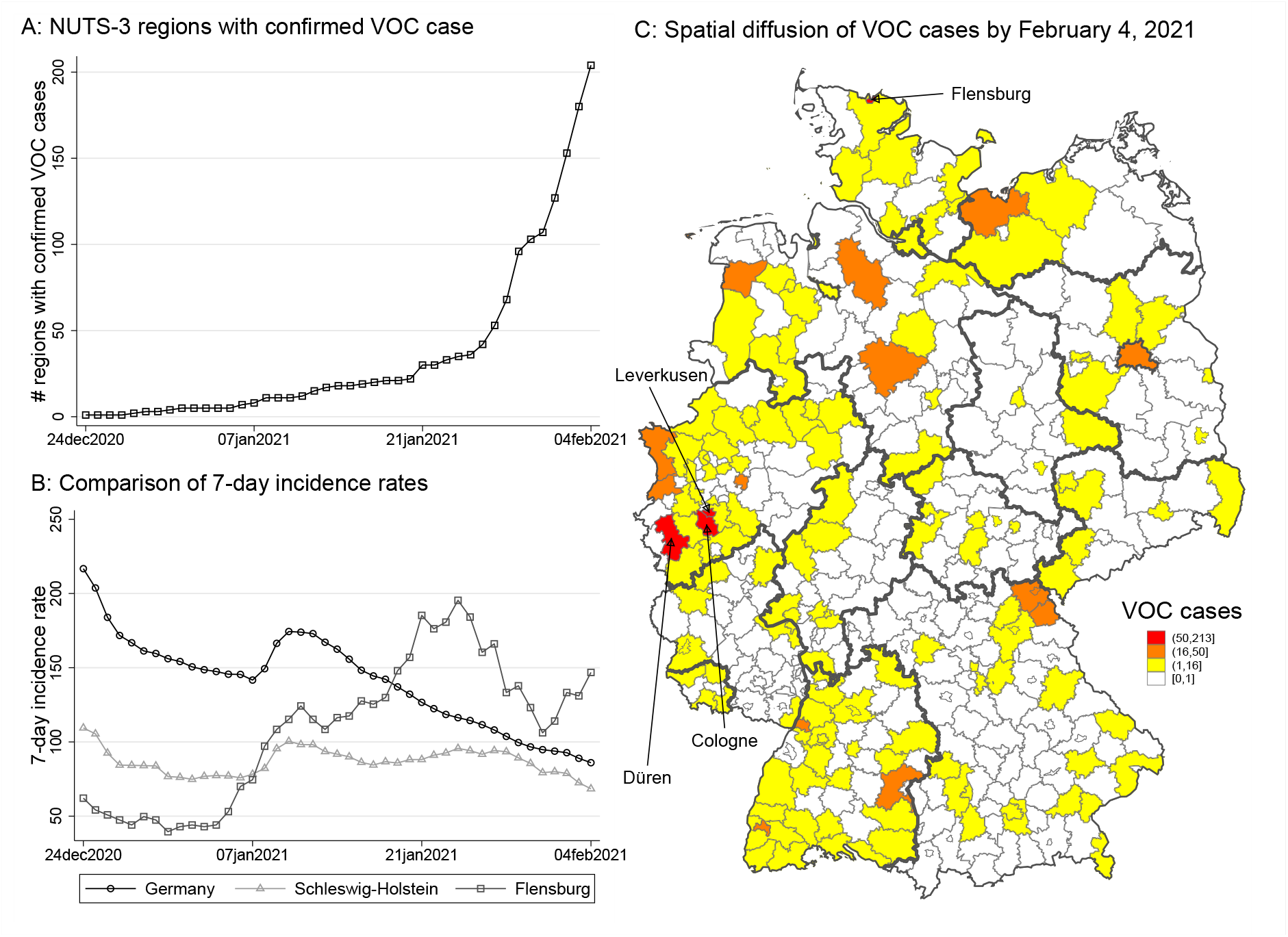
Temporal and spatial distribution of VOC cases in Germany. Panel A counts NUTS-3 regions with confirmed VOC cases over time. Panel B compares the 7-day incidence rate (by day of reporting) for Germany, Schleswig-Holstein, and Flensburg. Panel C shows the spatial spread of VOC cases by February 4, 2021. Flensburg and Cologne, Düren and Leverkusen report most confirmed cases.

## Results

### Synthetic Control Method (SCM)

In Figure 2, we estimate the change in incidence and hospitalization rates for Flensburg (Panel A and B) and the cluster of three NRW cities (Panel C and D) around the timing of the first VOC reporting. The figure displays the estimated percentage effect on these rates relative to their last pre-treatment values together with 90% confidence intervals (calculated from pseudo P values) to assess the statistical significance of the estimated changes in incidence rates. We set the start of the treatment period in the SCM analyses to January 5, 2021, which is at least one week before the confirmation of the first VOC cases in all four treated regions. Because we use infection data recorded by symptom onset, we argue that starting the treatment period at least one week before the first confirmed VOC case is sufficient to account for incubation times [9]. Hospitalization rates have previously been estimated to lag the infection development (onset of symptoms) by about 4-8 days [10].

**Figure 2.**
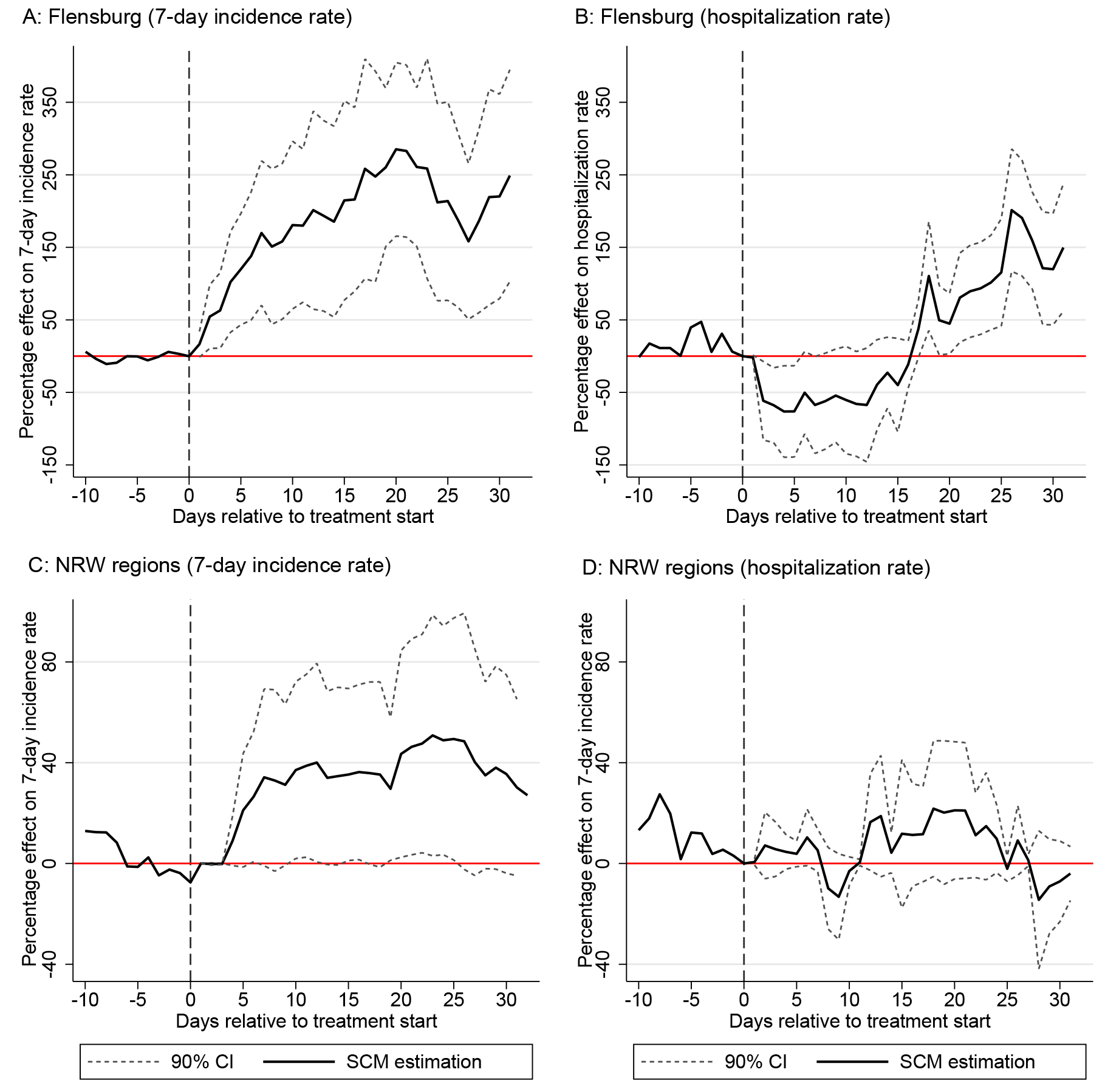
SCM estimates for the relative percentage increase in the 7-day incidence rate for Flensburg (Panel A) and NRW regions (Panel B) and the hospitalization rate for Flensburg (Panel C) and NRW regions (Panel D) vis-à-vis their synthetic control groups. Treatment start is set to January 5, 2021 in all cases. 90% confidence intervals are constructed on the basis of pseudo P values (see Method section for details).

Evaluated against the counterfactual infection development in the synthetic control group, the observed 7-day incidence rate for Flensburg becomes significantly larger after treatment start (see Panel A of Figure 2). No significant differences between treated and the synthetic control group are observed for the time period prior to treatment start. With regard to effect size, VOC spreading through illegal parties in Flensburg is associated with a tripling of the incidence rate (20 days after treatment start) compared to the counterfactual situation.

Consistent with prior evidence on the delay between infection and hospitalization, the hospitalization rate for Flensburg shown in Panel C of Figure 2 shows a significant increase approx. 16 days after treatment start. Although the estimated percentage change shows a doubling to tripling of hospitalized patients in intensive care 30 days after treatment start, we need to consider that this effect is calculated on the basis of a relatively small absolute number of hospitalized patients per 100,000 population at the local area level (i.e., 2 patients per 100,000 population in the last pre-treatment observation and, on average, 8 additional cases per 100,000 population in Flensburg during the treatment period).

The SCM results for the NRW cluster furthermore indicate that Flensburg may be a very exceptional case since illegal parties have likely served as super-spreading events for VOC infections. Treatment effect estimates for the NRW cluster report an increase in the 7-day incidence rate by approx. 40% (after 20 days) and no significant increase in hospitalization rates.

#### Difference-in-Difference (DiD) estimation

To comprehensively assess the complex correlation be-tween VOC cases and the 7-day incidence rate development across German regions, Table 1 reports results from difference-in-difference estimations, which compare the 7-day incidence rates in regions with and without confirmed VOC cases before and after VOC reporting. To increase estimation power, data on the three VOC types have been pooled. In Table 1 we report results for different time windows: in the baseline specification, we set the start to the date of the first VOC reporting in the region; we then extend the treatment period to 7, 14, and 21 days before this date to capture latent transmission from an imported VOC case.

**Table 1.** Difference-in-difference estimations for association between first reporting of variants of con-cern (VOC) and 7-day incidence rates at the local area level in Germany.

|  | (1) | (2) | (3) | (4) |
| --- | --- | --- | --- | --- |
| Treatment start | Day first reporting | 7 days before | 14 days before | 21 days before |
| **Panel A: Baseline** |  |  |  |  |
| VOC Reporting | 13.17**<br>[4.2,22.1] | 7.63<br>[-1.7,16.9] | 3.61<br>[-5.7,12.9] | 0.08<br>[-9.0,9.2] |
| N | 22441 | 22441 | 22441 | 22441 |
| NUTS-3 regions | 401 | 401 | 401 | 401 |
| **Panel B: First VOC reported before January 22, 2021** |  |  |  |  |
| VOC Reporting | 15.09+<br>[-1.7,31.9] | 17.83*<br>[2.7,32.9] | 17.74*<br>[2.6,32.8] | 13.65+<br>[-1.1,28.4] |
| N | 12712 | 12712 | 12712 | 12712 |
| NUTS-3 regions | 227 | 227 | 227 | 227 |
| **Panel C: More than 9 VOC cases reported [90% percentile]** |  |  |  |  |
| VOC Reporting | 35.00**<br>[17.9,52.1] | 30.65**<br>[17.3,44.0] | 24.63**<br>[11.6,37.7] | 17.61**<br>[4.8,30.4] |
| N | 13552 | 13552 | 13552 | 13552 |
| NUTS-3 regions | 242 | 242 | 242 | 242 |
| *The following applies to all panels* |  |  |  |  |
| Fixed Effects & Controls | Yes | Yes | Yes | Yes |

**Table 1.** Descriptive statistics DiD and Event Study.

|  | Mean | Std. Dev. | Min. | Max. |
| --- | --- | --- | --- | --- |
| **Panel A: Baseline with 7-day incidence rates** |  |  |  |  |
| 7-day incidence rate | 147 | 85 | 8 | 714 |
| VOC Reporting | .07 | .26 | 0 | 1 |
| Reported SARS-CoV-2 cases within the previous seven days | 296 | 399 | 5 | 8042 |
| Spatially Lagged reported SARS-CoV-2 cases within the previous seven days | 108 | 199 | 0 | 2778 |
| Mean temperature at 2 m above ground in °C | 1.8 | 3.5 | -11 | 14 |
| Daily mobility change in relation to 2019 | -15 | 17 | -73 | 76 |
| N | 22441 |  |  |  |
| **Panel B: Panel B: Baseline with Hospitalization** |  |  |  |  |
| Number of COVID-19 patients in intensive care per capita times 100,000 | 6 | 6 | 0 | 68 |
| Number of COVID-19 patients in intensive care with artificial ventilation per capita times 100,000 | 3.2 | 3.5 | 0 | 34 |
| N | 22217 |  |  |  |

*Notes:* Observations are for each NUTS-3 region per day. We always consider the time period between November 15, 2020 and February 4, 2021. In Panel A, we take into account all 401 NUTS-3 regions in Germany. By February 4, 2021, 204 NUTS-3 regions reported a VOC case and 197 did not. In Panel B, we take into account 397 NUTS-3 regions in Germany, for which we have daily data on the number of patients in intensive care. By February 4, 2021, 201 NUTS-3 regions reported a VOC case and 196 did not. Some observations have been dropped because we lack information on some control variables (e.g., on Daily mobility change in relation to 2019 for December 4-7, 2020).

We find a significant epidemiological trend associated with VOC reporting at the local area level. For the overall sample covering all regions with at least one confirmed VOC case, the estimates in Panel A point to an average increase in the incidence rate by 13.14 [95%-CI: 4.2, 22.1] cases per 100,000 population or approx. 12% (evaluated against the average 7-day incidence rate in comparative regions of 111). While effects are of similar size for the sub-sample estimates limiting treated regions to those with a VOC case confirmed before January 22 in Panel B (i.e., early treated regions for which we observe at least 14 days of treatment), we find a remarkably higher effect size for the subsample with at least 9 confirmed VOC cases (i.e., regions belonging to the top 10 percentile of absolute VOC counts). For the latter, we find an increase in the incidence rate of 32% on average (35.01 [CI: 17.9, 52.1] additional cases per 100,000 population). Generally, the extension of treatment periods by 7, 14 and 21 days ahead of the first VOC results in smaller point estimates in all three panels (with declining statistical significance). This provides a first indication that infection trends appear to change around the date of first VOC reporting and not significantly before that date.

Table 2 contains estimates for the baseline specification by variant type. In column (1), we focus on regions in which the British variant was certainly reported but potentially also other variants. In column (2), we condition only on regions with a reported British variant but no other variant. Column (1) and (2) confirm that the level of our previous estimates may mainly be associated with the British variant. In column (3), we focus on regions with reporting of the South African variant (but potentially also other reported variants in these regions by February 4, 2021). We find an increase in the incidence rate of 25% on average (28 [CI: 9.7, 46.1] additional cases per 100,000 population). However, this result is driven by the simultaneous presence of multiple variants (resulting in a higher VOC count as shown in Panel C of Table 1) because regions for which only the South African has been reported do not face higher incidence rates (see column (4)). We conclude that at this initial stage of VOC spread in Germany we cannot provide reliable estimates on individual VOC types but only for their joint statistical association with incidence rates at the local area level.

**Table 2.** Difference-in-difference estimations for association between first reporting of variants of concern type and 7-day incidence rates at the local area level in Germany.

|  | (1) | (2) | (3) | (4) |
| --- | --- | --- | --- | --- |
| Variant type: | With British | Only British | With South African | Only South African |
| VOC Reporting | 12.89**<br>[3.4,22.4] | 13.00<sup>+</sup><br>[-0.3,26.3] | 27.91**<br>[9.7,46.1] | -3.82<br>[-21.3,13.7] |
| N | 20593 | 18857 | 12488 | 11480 |
| NUTS-3 regions | 368 | 337 | 223 | 205 |
| Fixed Effects & Controls | Yes | Yes | Yes | Yes |

One major concern is that the estimated correlations between VOC reporting and the increase in the 7-day incidence rate as shown in Table 1 and Table 2 are simply a reflex of higher testing intensities in regions with confirmed VOC cases, which may then raise the number detected but potentially asymptomatic SARS-CoV-2 infections. To rule out this effect, we report DiD estimations results for the hospitalization rate as outcome variable in Table 3. We show estimates for the baseline specifications in analogy to Table 1 with treatment start being set to the day of first VOC reporting in a region.

**Table 3.** Difference-in-difference estimations for association between first reporting of variants of concern and number of COVID-19 patients in intensive care at the local area level in Germany.

|  | (1)<br>Baseline | (2)<br>First VOC reported<br>before Jan22, 2021 | (3)<br>With 9 or more VOC cases<br>reported [90% percentile] |
| --- | --- | --- | --- |
| **Panel A: All patients in intensive care** |  |  |  |
| VOC Reporting | 0.37<br>[-0.1,0.9] | 0.50<br>[-0.5,1.5] | 1.30**<br>[0.5,2.1] |
| N | 22217 | 12656 | 13496 |
| NUTS-3 regions | 397 | 226 | 241 |
| **Panel B: Patients with artificial ventilation** |  |  |  |
| VOC Reporting | 0.14<br>[-0.2,0.5] | -0.05<br>[-0.8,0.7] | 0.71**<br>[0.2,1.2] |
| N | 22217 | 12656 | 13496 |
| NUTS-3 regions | 397 | 226 | 241 |
| *The following applies to all panels* |  |  |  |
| Fixed Effects & Controls | Yes | Yes | Yes |

While the DiD results point to positive but insignificant correlations for the sample of all regions in column (1) and the subsample of regions with an early VOC reporting before January 22 in column (2), we find a statistically significant increase in the hospitalization rate for regions in the top 10% percentile of reported VOC cases in column (3). The estimated increase of 1.29 [CI: 0.5, 2.1] additional COVID-19 patients in intensive care per 100,000 population translates into a 42% rise compared to the average hospitalization rate in comparison regions (3.08 patients in intensive care per 100,000 population. Panel B illustrates that the results are robust to using the number of COVID-19 patients in intensive care with artificial ventilation per 100,000 population as the dependent variable.

### Panel Event Study (PES)

We finally employ a panel event study to investigate the time heterogeneity of the estimated correlations. Figure 3 reports daily estimates for the 7-day incidence rate (Panel A and B) and the hospitalization rate (Panel C and D) for the overall sample of regions and the subsample of treated regions belonging to the top 10% percentile of VOC cases. Effects are calculated for the relative time before and after VOC reporting in each region. Panel A shows for the sample of all regions with at least one VOC case that 20 days after VOC reporting the 7-day incidence rate has increased by approx. 48% compared to the average incidence rate in treated regions and the last pre-treatment observation. Panel A indicates 50 additional SARS-CoV-2 infections 20 days after treatment start compared to 105 infections per 100,000 population in the last pre-treatment period.

**Figure 3.**
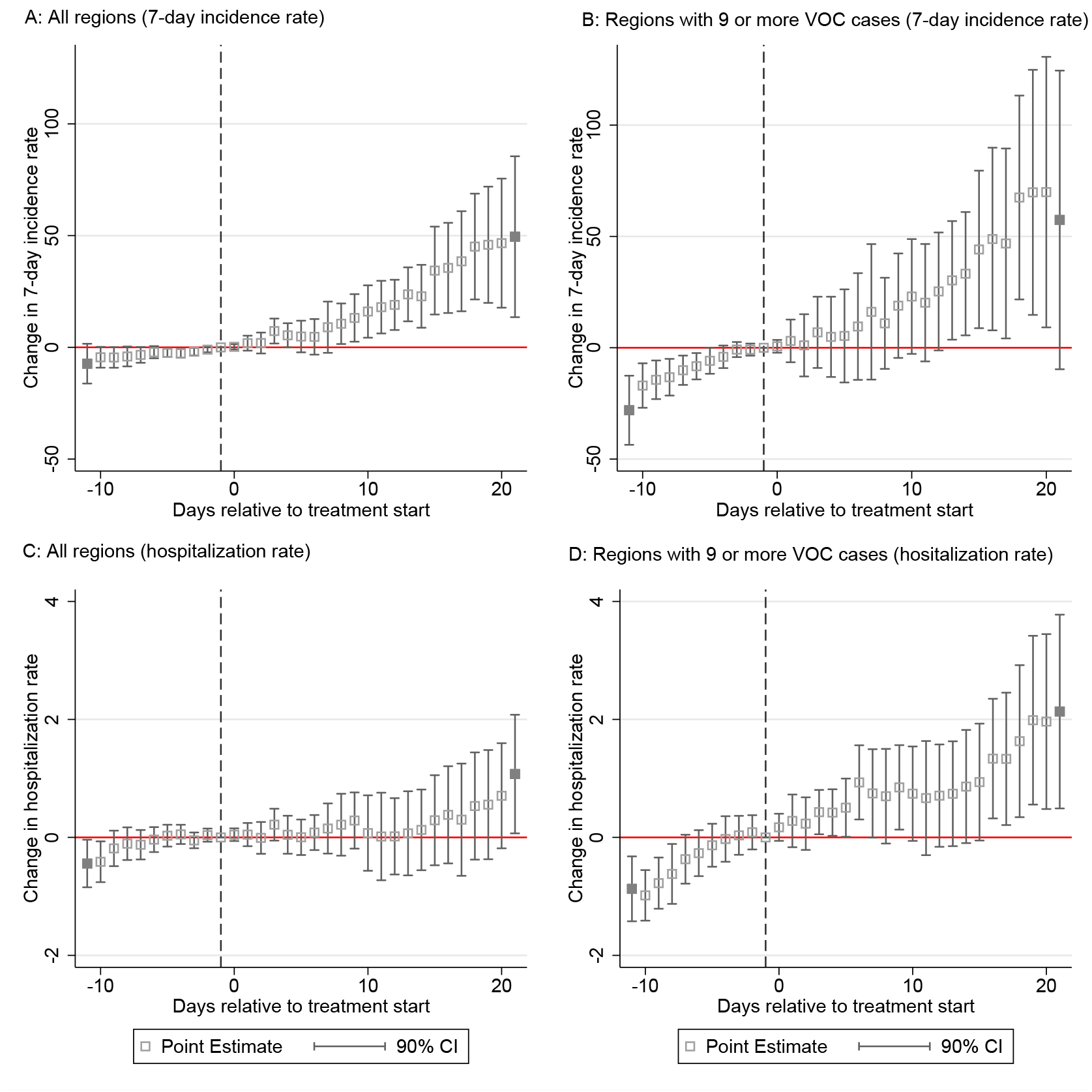
Panel event study estimates for trend development in the 7-day incidence rate (Panel A and B) and the hospitalization rate (Panel C and D) in regions with confirmed VOC cases around the day of first VOC reporting. Hollow squares show estimated daily point estimates; grey squares report cumulative estimates beyond the maximum number of leads (10) and lags (20) around the start of the first VOC reporting in treated regions. The dashed vertical line indicates that the last day before treatment start (−1) serves as benchmark period for the estimated daily lead and lag coefficients.

For regions in the top 10% percentile of VOC cases we find in Panel B of Figure 3 that the cumulative effect grows even stronger during this 20-day time window regional effect heterogeneity increases as well (as indicated by the widened confidence intervals). The fact that effects turn statistically insignificant beyond the 20-day time window may hint at mitigating effects from of strict containment measures implemented in some of these regions.

With regard to time dynamic effects for the hospitalization rate, significant effects can be observed approx. 15 days after treatment start for the sample of treated regions with 9 or more reported VOC cases. After 20 days, the hospitalization rate is estimated to be approx. 38% higher in treated regions compared to their last pre-treatment observation. This corresponds with two additional patients in intensive care per 100,000 in treated regions evaluated against the average of 5.19 patients in intensive care per 100,000 population before treatment start. Generally, Panel A to Panel D in Figure 3 clearly indicate that incidence and hospitalization rates in regions with VOC tend to grow over time relative to the counterfactual situation. The effects visualized in Figure 3 do not give indication for positive early anticipation effects from latent confounding events in treated regions determining the observe trend development. Our analysis thus indicates that the growth in incidence and hospitalization rates strongly coincides with the reporting of VOC cases at the local area level.

## Discussion

The main goals of this early empirical assessment were, firstly, to provide a near-time analysis of the initial dynamics of VOC spread within a country after international importation and, secondly, to estimate epidemiological trends associated with VOC reporting at the local area level in Germany. On average, we find that the 7-day incidence rate increased by around 30% in regions with confirmed VOC cases compared to regions without such a case after the first reporting. Moreover, a higher number of reported VOC cases at the local area is found to further increase the incidence rate in tendency.

For the comparative case study of Flensburg estimated by the synthetic control method, we even find a doubling to tripling of the incidence rate after first VOC reporting. This very large effect may, however, be the result of illegal parties operating as super-spreading events. The panel event study underlines a trend of growing infection rates in regions with VOC over time. This development is particularly alarming because we conduct our analysis for a time period with a relatively low overall share of VOC cases in Germany (around 6% between January 22 and January 29, 2021 [7]). Furthermore, the existence of latent VOC cases in comparison regions may lead to an underestimation of the correlations presented here.

Our results are limited in several important dimensions. A first complicating factor is that we cannot control for SARS-CoV-2 test intensities at the regional level. Test intensity may be higher in regions with confirmed VOC relative to comparison regions. Our results may partly capture this effect if more testing leads to a higher number of detected SARS-CoV-2 infections with only mild or no symptoms. To preclude a bias in the estimation results, we have analyzed the development of hospitalization rates at the local area level next to the 7-day incidence rate. Our findings also confirm a significant correlation between VOC reporting and the number of hospitalized patients per 100,000 population in those regions that have a relatively high number of VOC cases (top 10% percentile). We find that VOC importation to these regions is associated with an increase in the hospitalization rate by 40%. Also, the number of patients with artificial ventilation rises. This points to future threats for local health systems in regions with a high VOC spread.

Other limitations of our study should be addressed in future work: First, given the initial stage of VOC spread in Germany, we have mostly focused on a pooled data assessment across VOC types. While this approach increases estimation power, we can only identify differences in the correlation between local incidence rates and specific VOC types to a very limited extent. This is further complicated by the simultaneous presence of different variants in several regions. Second, we only know the day of reporting of a VOC case but not the *de facto* arrival of virus mutations in a region, which makes estimation difficult.

Third, we only estimate the correlation between confirmed VOC cases and the 7-day incidence rate of infections or the hospitalization rate. While our results provide robust evidence against the statistical null hypothesis of no association between the presence of VOC and the development of local infection and hospitalization rates, we cannot infer the causal nature of this relationship. I.e., our results indicate but do not prove whether VOC have a higher transmissibility compared to previously existing SARS-CoV-2 strains or not. Representative genomic data at the local area level is needed to comprehensively answer this question [11]. Until such data are available, we hope that our early assessment can fill the eminent knowledge gap on this matter and inform public health authorities about the need for actions to mitigate global and local transmission at this initial stage [12].

## Methods

We employ complementary statistical tools to systematically estimate the complex correlation structure between VOC reporting, local incidence rates and hospitalization rates. Given the unequal spatial distribution of confirmed cases and the emergence of local clusters with relatively high VOC counts (as shown in Panel C of Figure 1), we start out with comparative case studies for most severely affected regions, namely Flensburg, on the one hand, and the cluster of three NUTS-3 regions (Cologne, Leverkusen and Düren), on the other hand. Estimations are conducted by the synthetic control method (SCM) for single and multiple treated units, which compares the infection development in the case study regions to the development in a synthetic control group composed of similar regions without confirmed VOC cases [13, 14].

To establish a meaningful counterfactual situation, the SCM approach constructs the synthetic control group as the weighted average of regions in the donor pool, i.e. all regions without reported VOC case. Weights are estimated through a minimum distance approach for predictor values in the pre-treatment period. We provide an overview of predictors used for SCM estimation together with a description of SCM implementation and statistical inference in the SI appendix. While SCM offers favorable features for detailed case study analyses related to COVID-19 research [15, 16, 17], its focus on selected regions limits its ability to provide a comprehensive picture of the epidemiological trends associated with reporting of VOC cases across all German NUTS-3 regions.

We therefore complement SCM estimation by a systematical assessment of this link using difference-in-difference (DiD) regressions and a panel event study (PES). The main idea of the DiD approach is to identify differences in epidemiological trends (time differences) across regions with and without treatment (cross-sectional difference), where treatment is defined as the reporting of at least one VOC case at the regional level. We also use alterations in the definition of the treatment group, for instance, to investigate if the strength of the association between VOC reporting and the development of epidemiological indicators changes with the relative intensity of local VOC spread. A review of the suitability of the DiD approach for COVID-related empirical research is given in [18].

The complementary PES approach finally seeks to investigate time heterogeneity in the estimated correlations that may stem from a stronger correlation between treatment and outcome variables over time [19]. One attractive feature of PES is that it can properly handle staggered treatment start, i.e., in our case the first reporting of a VOC case at the local area level. Different from the DiD approach, which estimates average treatment effects in absolute time by calendar days, the PES measures time for each treated region relative to treatment start. This increase the robustness of estimates around the timing of treatment start. Given the dynamic nature of COVID-related epidemiological data and the staggered nature of events, e.g. policy actions to mitigate disease spread, PES has been frequently applied in several COVID-related studies [20, 21, 22].

We provide an extended method section in the SI Appendix including a description of identification challenges and how we approach them. To ensure comparison across estimation methods, we apply all three approaches to the same set of outcome variables, i.e. the 7-day incidence rate and the number of hospitalized patients in intensive care (with and without artificial ventilation) per 100,000 population. Data on daily SARS-CoV-2 infection data (by the onset of symptoms) for each of the 401 NUTS-3 regions has been collected from the COVID-19 dashboard of the Robert Koch Institute (RKI), which is in charge for disease surveillance in Germany [23]. The number of hospitalized patients in intensive care for 397 NUTS-3 regions with an own hospital is taken from the INFAS corona database [24].

We account for several confounding factors in the link between the treatment indicator and the evolution of the outcome variable. These include daily mobility patterns and daily weather data at the regional level together with time-invariant demographic structures of regions. The link between seasonality and infection dynamics has recently been studied [25]. Including mobility effectively controls for social interaction as a driver of local infection dynamics and also for lockdown measures implemented during the sample period (and how closely people followed these rules) [26]. Given that we use finely granulated regional data, we also account for spatial spillovers in local SARS-CoV-2 infection numbers by calculating spatial lag variables capturing the SARS-CoV-2 development in the geographical neighborhood of each NUTS-3 regions. The SI appendix contains data descriptions and source information.

As default specification, we pool information on VOC reporting over individual variant types. This shall increase estimation power given that the absolute number of reported VOC cases during our sample period is still low. As a robustness check we further provide tentative estimates by individual VOC types based on the available information on genomic sequencing in the VOC event database. It is further important to stress that data in the VOC event database rely on public reporting in newspapers and other publicly available documents and webpages. While we have cross-checked data consistency of VOC reporting by retracing individual cases and their timing from the documented sources in [8], unfortunately, we cannot make statements about the completeness of the data on VOC cases.

The only available alternative VOC data at the local area level, which are not publicly available yet, are the results of a joint reporting system of the Robert Koch Institute (RKI), which is charge for disease surveillance and control in Germany and local health authorities. According to a monitoring report by the RKI [7], this joint reporting system had registered VOC cases in 13 out of 16 federal states by end of January. In comparison, the VOC event database used here had already identified confirmed VOC cases in all 16 federal states by that date. We therefore argue that –in the absence of alternative data sources at the local area level– the empirical approach taken here offers a feasible way for providing near-time information on VOC spread in Germany and to assess the statistical association between VOC spread and epidemiological trends.

Nonetheless, in the light of the mentioned data uncertainties, it is of paramount importance that the results presented here need to be interpreted carefully: First, because information is limited to the day of VOC reporting but not the *de facto* arrival of virus mutations in a region. Second, in the international comparison, VOC testing rates are still relatively low in Germany and we thus do not know the latent degree of VOC diffusion at the regional level. Third, using data at the population level for NUTS-3 regions only allows us to identify correlations between VOC reporting and epidemiological trends, but no causal statements can be made whether VOC have a higher transmissibility than previously circulating virus strains and whether VOC infections lead to more severe COVID-19 disease cases. Such statements would require alternative data, ideally based on comprehensive contact tracing data for individual infections with different SARS-CoV-2 strains [27].

## Data Availability

Data is public. Replication files will be available with published version.

## Data availability

All study data and codes to replicate the estimation results (including robustness tests) are stored in a publicly available data repository (will be available with published version).

## Author contributions

Both authors contributed equally to this article and share first authorship; both performed the analysis, acquired data, wrote and edited the manuscript.

## Supplementary Information for

### Extended Methods

#### Synthetic control method (SCM)

We use SCM to analyse two case studies for a single treated unit (Flensburg) and multiple treated units (cluster of 3 cities/regions [Cologne, Leverkusen and Düren] in North Rhine-Westphalia). In all four case study regions a SARS-CoV-2 variant of concern (VOC) has been confirmed by genome sequencing, which is used to identify treatment status. VOC are defined as SARS-CoV-2 mutations from the British B.1.1.7, the South African B.1.351 and the Brazilian P.1 variants. The objective of SCM is to compare the development in epidemiological outcome variables after treatment start in the two sets of treated region(s) vis-à-vis a synthetic control groups selected from a donor pool of 197 comparison regions without any confirmed VOC case during the entire sample period December 15, 2020 to February 4, 2021. Outcome variables are i) the 7-day incidence rate (SARS-CoV-2 infections per 100,000 population over the last seven days) and ii) the hospitalization rate (hospitalized patients in intensive care per 100,000 population). We compute daily treatment effects for a maximum of 31 days and express them as percentage difference to the last pre-treatment observation of the scaled outcome variable (to 100), see Fig. 2 in the main text.

We motivate the use of SCM as one element of our empirical identification strategy because the estimation approach has been shown to be a flexible and robust estimation tool that has previously been applied to COVID-related research, for instance, to study the effect of face masks on SAR-CoV-2 infection numbers in Germany [1] and lockdown effectiveness for a counterfactual of Sweden [2] and the USA [3]. The key identification approach of SCM is to establish a counterfactual that mimics a situation in which the treatment in treated regions (here: the emergence of VOC cases) would not have taken place. This is implemented by means of creating a synthetic control group consisting of the donor pool of comparison regions and by comparing the outcomes of treated units and the synthetic control after the start of the treatment. The match between treated regions and the synthetic control group is done through a minimum distance approach for a set of predictor variables evaluated along their pre-treatment values for treated regions and those in the donor pool. This ensures that pre-treatment differences in trends of the outcome variable are leveled. A formal description of the estimation approach and the underlying assumptions for effect identification are given in [4, 5, 6].

For our purpose of estimating the epidemiological effects of emerging VOC in German regions, we adopt and extend the data and estimation setup applied in [1]. For both SCM applications, we set the start of the treatment period to January 5, 2021 and identify treatment effects of VOC throughout January and early February. In all four cases, the reporting of the first confirmed VOC case took place at least one week after the start of the treatment period (Cologne: January 12, 2021, Leverkusen: January 18, 2021, Düren: January 23, 2021, Flensburg: January 24, 2021). This time lag between the start of the treatment and the reporting of the first VOC case should ensure that latent transmission effects are captured in the estimation. For instance, in the case of Flensburg, VOC infections could be traced back to illegal parties on December 31, 2020 [7]. Considering a median incubation time of 5 days for SARS-CoV-2 infections [8], we can thus expect that first VOC effects become visible in the data from January 5, 2021 onwards. This is before the first VOC case was confirmed through genome sequencing on January 24, 2021 for Flensburg.

Data on reported SARS-CoV-2 infections are taken from the Robert Koch Institute [9]. For our empirical analysis we use aggregate case numbers for each NUTS-3 region and day tracked on the basis of symptom onset for individual cases rather than the reporting date by local health authorities. This allows us to estimate the transmission timing of SARS-CoV-2 at the regional population level more precisely. We aggregate the data across age groups. Data on confirmed cases of the three novel VOC (B.1.1.7., B.1.351, P.1) together with their reporting dates are gathered from a public crowd-sourcing project [10], which bases case documentation on newspaper and public health reports. We have cross-checked data consistency by retracing individual cases and their timing from the documented source information and have conducted additional online searches for selected cases.

A relevant concern against SARS-CoV-2 case numbers is that they may grow with growing test intensity. Test intensity may rise in regions in regions with confirmed VOC cases. To rule out this effect, we use the hospitalization rate as alternative outcome. Specifically, we use daily information on the number of COVID-19 patients in intensive care (with and without artificial ventilation) per 100,000 population [11].

In the specification of SCM estimation, we account for the autoregressive dynamics of infections and the hospitalization rate by including the 7-day incidence rate and the absolute number of cumulative SARS-CoV-2 infections during the last 3 weeks before treatment start as time-varying predictors. Other time-varying predictors are the average daily temperature for each region during the last 2 weeks and changes in average daily mobility during the last 2 weeks before treatment start. Changes in average daily mobility per region are measured relative to a 2019 (pre-COVID) benchmark period. We use data on daily temperatures from Deutscher Wetterdienst [12] and data on mobility changes from [13].

We further include time-constant cross-sectional predictors characterizing regional demographic structures and the regional health care system as in [1] based on data from the INKAR online database of the Federal Institute for Research on Building, Urban Affairs and Spatial Development [14]. We use the latest year available in the database, which is 2017. Employed cross-sectional predictor variables include population density (Population/km2), regional settlement structure (categorial dummy), the share of highly educated population (in %), the share of female in population (in %), the average age of female and male population (in years), old- and young-age dependency ratios (in %), the number of physicians per 10,000 of population and pharmacies per 100,000 of population.

We conduct all SCM estimations in STATA using the SYNTH [15] and SYNTH_RUNNER [16] packages. Confidence intervals (CIs) are calculated from one-sided pseudo *p*-values obtained on the basis of comprehensive placebo-in-space tests. The latter tests calculate pseudo-treatment effects for all regions in the donor pool treating each of the regions as if it would have received the treatment of a confirmed VOC case by or after January 5, 2021. One-sided pseudo *p*-values are then calculated of the share of placebo-treatment effects that are larger than the observed treatment effects for treated regions and thus indicate the probability that the increase in the number of SARS-CoV-2 infections was observed by chance given the distribution of pseudo-treatment effects in the donor pool. To account for differences in pre-treatment match quality of the pseudo-treatment effects, only donors with a good fit in the pre-treatment period are considered for inference. Specifically, we do not include placebo effects in the pool for inference if the match quality of the control region, measured in terms of the pre-treatment root mean squared prediction error (RMSPE), is greater than 10 times the match quality of the treated unit [6]. Based on the obtained pseudo *p*-values we calculate confidence intervals as described in [17].

##### Robustness

We mainly perform robustness tests by changing the composition of the donor pool. First, we exclude donor regions that were selected in the baseline SCM estimation. The idea behind this analysis is to preclude unintended selection effects resulting from latent VOC transmissions captured in the overall infection dynamics of donor regions in the pre-treatment period. Second, we reduce the pool of donor regions to those NUTS-3 regions which are located in the same federal state as the treated regions (Schleswig-Holstein for Flensburg and North Rhine-Westphalia for Cologne, Leverkusen and Düren). This approach should minimize differences in public health measures, which are mainly decided under the authority of individual federal states in Germany. While mentioned but not reported in the manuscript in detail, we include codes to run all robustness tests in the replication files.

#### Difference-in-difference estimation (DiD)

To investigate average and dynamic treatment effects for the entire group of treated regions with at least one confirmed VOC case, we additionally run a series of panel regressions in a DiD and Panel event study (PES) framework. The sample period for the panel regressions includes the time period between November 15, 2020 and February 4, 2021. This ensures that we cover all confirmed VOC cases (in the currently best possible way) together with a sufficient pre-treatment period for each region of at least 3 weeks. As for the case of SCM, we use the 7-day incidence rate and the hospitalization rate as key outcome variables. For the incidence rate we measure the timing of infections in terms of symptom onset rather than reporting by local health authorities. We also use the same set of time-varying predictor variables as described above; cross-sectional predictors for the set of NUTS-3 regions are not included as we account for NUTS-3 region fixed effects in the panel regressions.

Specifically, we run DiD regressions as two-way fixed effects model of the following general form

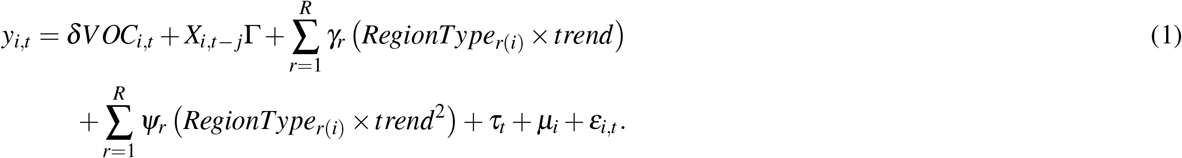

In equation 1, *y*_*i,t*_ is the epidemiological outcome of interest (7-day incidence rate, hospitalization rate) observed for NUTS-3 region *i* and day *t*. The variable *VOC*_*i,t*_ is our main treatment indicator, which takes values of 1 from the day onwards for which the first VOC case was confirmed in the region. The coefficient *δ* measures the direction and strength of the correlation between VOC reporting and outcome variables. We additionally test for latent transmission effects prior to the first reporting of a VOC (given that genome sequencing to identify mutations may take up to 2 weeks). This is done by moving forward the date when the treatment dummy *VOC*_*i,t*_ takes values of 1 for treated regions by 7, by 14 and by 21 days, respectively. Importantly, these extended treatment specifications do not test for early anticipation effects caused by latent confounding factors (this is done in the Panel Event Study), but averages estimated effects over a longer treatment period to capture potential latent transmissions prior to the first VOC confirmation as, for instance, identified for Flensburg in the SCM estimations.

It is important to control for factors that potentially confound the link between VOC and the overall SARS-CoV-2 incidence rate at the regional level. The set of confounding factors, which we can directly account for, is included in the variable vector *X*_*i,t− j*_. Specifically, similar to the SCM application, we control for the number of SARS-CoV-2 cases in region *i* during the last, the second last and the third last week. We also include a spatially lagged variable covering the number of SARS-CoV-2 cases in region *i*’s (direct) spatial neighbors during the last, the second last and the third last week. A spatial lag is important because infections can easily spread from one region to another region nearby, e.g., due to commuting or general mobility [18]. Spatial association between regions is measured through first-order contiguity, i.e. whether regions share a common border or not. We row standardize the resulting spatial weights matrix.

Further, we control for the average temperature [19] and the relative change in average daily mobility at *t—* 1, at *t−* 7 and at *t−* 14 in region *i*. Controlling for mobility is important because lower mobility can be disease mitigating [20, 21, 22]. Including mobility effectively controls for lockdown measures implemented during the sample period and how people follow the rules. In addition, we include linear and quadratic time trends for four different region types (*RegionType*_*r*(*i*)_) classified on the basis of the region’s settlement structure including region type 1 (large district-free cities, kreisfreie Städte), type 2 (urban regions, Landkreise), type 3 (rural regions, Landkreise) and type 4 (sparsely populated regions, Landkreise), i.e. *R* = 4. The classification of region types follows the definition of the Federal Institute for Research on Building, Urban Affairs and Spatial Development [14]. Table 1 shows descriptive statistics.

*τ*_*t*_ are time fixed effects for each day in the sample, which for instance control for daily changes in infection levels similar across regions. *µ*_*i*_ controls for time-constant region fixed effects, which could, e.g., be caused by region-specific SARS-CoV-2 testing intensities. *ε*_*i,t*_ is the model’s error term. We cluster standard errors at the NUTS-2 level (each of the 401 NUTS-3 regions belongs to one of the 38 NUTS-2 regions in Germany). We estimate *δ* and Γusing the REGHDFE package [23] in STATA, which allows us to control for *τ*_*t*_, *µ*_*i*_, *γ*_*r*_ and *Ψ*_*r*_.

##### Robustness

Besides the full sample covering all treated regions, we also conduct estimations with sub-samples. First, we focus on those experiencing treatment early on (before January 22, 2021) to observe at least 14 days of treatment after the first confirmed VOC case for each treated region. Second, we study regions with a relatively high number of VOC reported cases. Here, we restrict the treated regions to the top-10 percentile of VOC counts, which corresponds to at least 9 VOC cases per region. The idea of this subsample is to test for treatment effect difference associated with confirmed VOC counts rather than the presence of at least one VOC case. Finally, we run estimations by variant type. In a first sub-sample, we focus on the British variant but allow for other reported variants. In a second sub-sample, we only consider the British variant. Similarly, we investigate effects for regions where the South-African variant was reported (together with potential other variants) and for regions for which only the South African variant was reported.

#### Panel event study (PES)

The estimation of the PES differs from the two-way fixed effects DiD specification mainly in the way that it accounts for the staggered emergence of a VOC in treated regions throughout the sample period. This allows us to identify dynamic treatment effects over time. Dynamic treatment effects may arise for different reasons: First, they could reflect early anticipation effects prior to the treatment start due to latent VOC transmissions or, second, they could result from other unobserved confounding factors systematically affecting incidence rates in treated regions around the treatment start. Thus, it is important to test for such early anticipation effects. The absence of statistically significant estimates for the latter but significant treatment effects could, accordingly, be interpreted in favor of our empirical identification strategy.

Moreover, we may expect that infection and hospitalizations effects do not immediately occur after the reporting of the first VOC case but potentially build up over time at the regional population level. This may particularly be the case if public health authorities can only imperfectly trace and mitigate VOC-related disease spread. In this case, the estimation of average treatment effects on treated (ATTs) as shown in equation 1 may potentially underestimate the dynamics of SARS-CoV-2 infections and hospitalizations subject to confirmed VOC cases. By including sufficient lag and lead terms in the estimation framework for the timing of treatment start, we can identify dynamic treatment effects.

The staggered nature of treatment start in different treated regions can be incorporated into the panel regression approach by translating the model from a specification in absolute time *t* (as shown in equation 1) to a specification that measures time for each region relative to treatment start. Together with the recognition of potential heterogeneity in the strength of treatment effects over time, the PES setup allows us to precisely estimate the impact of the passage of a treatment (here: VOC reporting in a region) that occurs at different times in different spatial units. A more formal presentation of the PES approach together with estimation challenges is given in [24, 25] among others. Prior COVID-related PES applications have dealt, for instance, with the infection effects from school re-opening in Germany [26] and [27], university students traveling during the U.S. spring break [28] or mass protests from the Black Lives Matter movement [29].

In the implementation of the PES approach, we include the same set of covariates and fixed effects as in the case of the two-way fixed effects DiD estimation. We also cluster standard errors at the NUTS-2 level. We set the maximum number of pre-treatment leads to 10 days and the maximum number of lags after the treatment start to 20 days. Further effects from leads/lags before (after) this range are accumulated to a single coefficient. To allow for an easy comparison of the SCM and PES results, we express all reported effects as percentage change relative to the observed 7-day incidence rate in the last pre-treatment period.

##### Robustness

Besides the full sample covering all treated regions, we estimate the effects for sub-samples of regions. First, we focus on those experiencing treatment early on (before January 22, 2021 to observe at least 14 days of treatment after the first confirmed VOC case). Estimation results for this sub-sample are only shown in the replication files. Second, we study regions with a relatively high number of VOC reported cases. We restrict the treated regions to the top-10 percentile of VOC counts, which corresponds to at least 9 VOC cases per region.

We conduct the PES estimations in STATA. The analysis builds on the EVENTDD package [30]. We document the full analyses in the replication files, particularly the mentioned robustness tests.

